# Protection conferred by Delta and BA.1/BA.2 infection against BA.4/BA.5 infection and hospitalization: A Retrospective Cohort Study

**DOI:** 10.1101/2022.11.14.22282310

**Authors:** Nicole E Winchester, Nabin K. Shrestha, Priscilla Kim, Larisa G. Tereshchenko, Michael B Rothberg

## Abstract

**Background:** SARS-CoV-2 immunity has declined with subsequent waves and accrual of viral mutations. In vitro studies raise concern for immune escape by BA.4/BA.5, and a study in Qatar showed moderate protection, but these findings have yet to be reproduced.

**Methods:** This retrospective cohort study included individuals tested for COVID-19 by PCR during Delta or BA.1/BA.2 and retested during BA.4/BA.5. The preventable fraction (PF) was calculated as ratio of the infection/hospitalization rate for initially positive patients divided by infection/hospitalization rate for initially negative patients, stratified by age, and adjusted for age, gender, comorbidities, and vaccination using logistic regression.

**Results:** 20,987 patients met inclusion criteria. Prior Delta infection provided no protection against BA.4/BA.5 infection (Adjusted PF: 11.9% (95% confidence interval [CI], 0.8-21.8); p=0.036) and minimal protection against hospitalization (Adjusted PF: 10.7% (95%CI, 4.9-21.7); p=0.003). In adjusted models, prior BA.1/BA.2 infection provided 45.9% (95%CI, 36.2-54.1) (p <0.001) protection against BA.4/BA.5 reinfection and 18.8% (95% CI, 10.3-28.3) (p<0.0001) protection against hospitalization. Up-to-date vaccination provided modest protection against reinfection with BA.4/BA.5 and hospitalization.

**Conclusions:** Prior infection with BA.1/BA.2 and up-to-date vaccination provided modest protection against infection with BA.4/BA.5 and hospitalization, while prior Delta infection provided minimal protection against hospitalization, and no infection protection.

## Introduction

Since the emergence of SARS-CoV-2 in 2019, multiple new strains have emerged, challenging the protection afforded by natural immunity, vaccination, and neutralizing antibody treatments. In the United States, these variants have included B.1.1.7 (Alpha), B.1.351 (Beta), B.1.1.28.1 (Gamma) and B.1.617.2 (Delta) [1], and most recently Omicron [1]. Prior to the emergence of Omicron, previous infection appeared to provide 80-90% protection against reinfection [2-9]. However, infection with pre-Omicron variants only provided 18-69% protection against reinfection with Omicron [10-11]. Since then, additional spike protein mutations have accrued, enabling progressive immune escape by new Omicron sub lineages [12-14].

By June 25^th^, two new Omicron subvariants, BA.4 and BA.5, became dominant in the United States [1]. *In vitro* studies raise concern that their additional spike protein mutations may enable immune evasion and repeat Omicron infections. Antibodies generated against the spike protein by BA.1 infection are evaded by BA.4/BA.5 [12], and neutralizing antibody titers following triple vaccination [13] or previous infection with BA.1 or BA.2 are substantially lower [13]. However, these studies have been limited to *in vitro* analysis of humoral immunity. A recent study in Qatar showed that infection with pre-Omicron variants was 28% effective at protecting against BA.4/BA.5 infection, while infection with early Omicron variants was 76% effective [15]. However, these findings have yet to be reproduced or evaluated in other populations, including the US, where patients are substantially older [15]. Moreover, this study did not assess the impact of prior infection or vaccination on hospitalization. Clinical data are needed to define such protection in order to guide quarantine and isolation guidelines and future vaccine development.

As the pandemic has progressed, the public health priority has shifted from preventing all infections to reducing disease spread and disease severity, especially hospital admissions. We therefore designed our study to address two specific aims: 1) to determine the degree to which infection with Delta or Omicron subvariants BA.1/BA.2 and up-to-date vaccination protects against reinfection with BA.4/BA.5 and 2) to determine the preventable fraction of BA.4/BA.5 hospitalizations attributable to prior infection with Delta or BA.1/BA.2 and up-to-date vaccination.

## Methods

### Study Design

We conducted a retrospective cohort study of patients within the Cleveland Clinic health system.

### Study Populations and SARS-CoV-2 Exposures

The study population included individuals aged 18 years or older tested for COVID-19 via polymerase chain reaction (PCR) between July 1, 2021 and August 18, 2022. To assess the protection afforded by Delta and early Omicron (BA.1/BA.2) infection against BA.4/BA.5, the data were divided into two cohorts according to the CDC reported variant proportions. Patients tested between July 1, 2021 and December 25, 2021 were included in the Delta cohort. Patients tested between December 26, 2021 and June 24, 2021 were included in the BA.1/BA.2 cohort.

Only patients tested during Delta and/or BA.1/BA.2 and then retested during the period of BA.4/BA.5 predominance (June 25, 2022 to August 18, 2022) were included. Patients with a positive COVID test before July 1, 2021 or during both the Delta and BA.1/BA.2 waves were excluded. In addition, because the CDC defines reinfection as occurring > 90 days after initial testing, patients with a positive test within 90 days of BA.4/BA.5 testing were also excluded. Patients with PCR testing during Delta and BA.1/BA.2 were included in both analyses if they only tested positive during one wave, or neither. The reasons for PCR testing included symptoms consistent with COVID-19, hospital admission for any indication, pre-procedural screening, and travel clearance.

COVID vaccination status at the time of BA.4/BA.5 predominance was confirmed by review of the EMR. Patients were classified as up-to-date if they had completed a two-dose series (Moderna or Pfizer) or a single dose vaccine (Johnson & Johnson’s Janssen or Astra Zeneca vaccine) and then received an additional vaccine dose within six months prior to June 25, 2022.

### Data Collection

Data were extracted from the electronic medical record. Variables collected included age, sex, BMI, ICD-9 codes for hypertension, diabetes mellitus, chronic kidney disease, heart failure, and stroke as well as COVID vaccine dose type and date(s), COVID PCR test date(s) and indications, hospitalization, ICU admission, and mechanical ventilation.

### Statistical Analysis

Infection rates were calculated as the number of positive tests divided by the total number of subjects retested. For aim 1, the fraction of BA.4/BA.5 infections that could have been prevented (preventable fraction (PF)) by prior infection with Delta or BA.1/BA.2 was calculated as 1 minus the ratio of the infection rate for initially positive patients divided by the infection rate for initially negative patients.

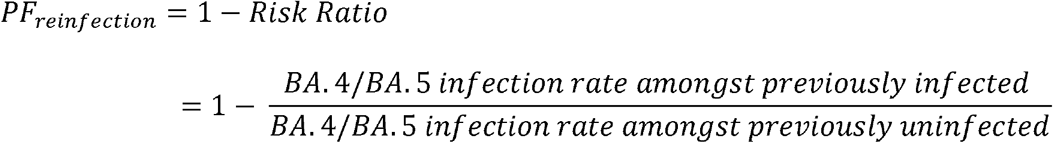

To assess for differences in protection by age, we stratified the preventable fraction by age (18-64 years, 65 years and up). Lastly, the adjusted PF of infection was calculated by logistic regression, comparing the odds of having versus not having prior SARS-CoV-2 infection among patients with BA.4/BA.5 infection as:

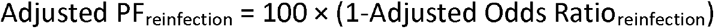

Logistic regression models were adjusted for age, sex, body mass index (BMI), and comorbidities (hypertension, diabetes mellitus, chronic kidney disease, heart failure, and stroke). Each model included an indicator variable for prior infection during Delta or BA.1/BA.2 and an indicator variable for up-to-date vaccination.

For the second aim, the adjusted PF was calculated for hospitalization. The fraction of BA.4/BA.5 hospital admissions that could have been prevented (PF) by prior infection with Delta or BA.1/BA.2 was calculated as:

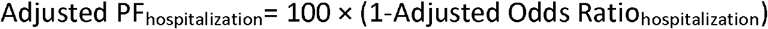

Logistic regression models were adjusted for age, sex, BMI, comorbidities (hypertension, diabetes mellitus, chronic kidney disease, heart failure, and stroke), and included an indicator variable for prior infection during Delta or BA.1/BA.2 and up-to-date vaccination.

Analyses were conducted using R v.4.2.1 (R Core Team, Vienna) and STATA/MP 17.0 (StataCorp, College Station, TX). To adjust for testing of 10 hypotheses, p-value < 0.005 was considered to be statistically significant. The Cleveland Clinic Institutional Review Board approved this work.

## Results

Between July 1, 2021 and August 18, 2022, 20,987 individuals met study inclusion criteria (average age, 58.5 ± 19.2 years; 57.4% female). Characteristics of patients who received a PCR test during the period of Delta, BA.1/BA.2, and BA.4/BA.5 predominance are shown in Table 1. During the Delta wave, 15,658 patients were tested for SARS-CoV-2 by PCR, of whom 15.2% tested positive. During the period of Omicron subvariants BA.1/BA.2 predominance, 10,545 patients were tested for SARS-CoV-2 by PCR, of whom 17.9% tested positive. Overall, 17.0% of these individuals retested positive during BA.4/BA.5.

**Table 1.** Patient characteristics and unadjusted preventable fractions (PF) for prevention of BA.4/BA.5 infection by prior SARS-CoV-2 infection status

| Characteristic | Delta |  | BA.1/BA.2 |  | BA.4/BA.5 |  |
| --- | --- | --- | --- | --- | --- | --- |
|  | SARS-CoV-2(+) | SARS-CoV-2(-) | SARS-CoV-2(+) | SARS-CoV-2(-) | SARS-CoV-2(+) | SARS-CoV-2(-) |
| n | 2,380 | 13,278 | 1,886 | 8,659 | 3,558 | 17,429 |
| Age ± SD, y | 52.3 ± 19.1 | 58.9 ± 19.1 | 55.7 ± 19.5 | 60.7 ± 18.4 | 56.4 ± 19.4 | 59.0 ± 19.1 |
| Female, n (%) | 1,378 (57.9) | 7,785 (58.6) | 1,075 (57.0) | 4,781 (55.2) | 2,093 (58.8) | 9,956 (57.1) |
| HTN, n (%) | 1,210 (50.8) | 7,976 (60.1) | 1,066 (56.5) | 5,597 (64.6) | 1,857 (52.2) | 10,539 (60.5) |
| DM, n (%) | 597 (25.1) | 3,902 (29.4) | 539 (28.6) | 2,781 (32.1) | 846 (23.8) | 5,112 (29.3) |
| CHF, n (%) | 361 (15.2) | 2,885 (21.7) | 373 (19.8) | 2,183 (25.2) | 505 (14.2) | 3,799 (21.8) |
| CKD, n (%) | 363 (15.3) | 2,816 (21.2) | 388 (20.6) | 2,120 (24.5) | 566 (15.9) | 3,701 (21.2) |
| Stroke, n (%) | 347 (14.6) | 2,824 (21.3) | 370 (20.6) | 1,971 (22.8) | 556 (15.6) | 3,666 (21.0) |
| Overweight, n (%) | 623 (26.2) | 3,828 (28.8) | 506 (26.8) | 2,476 (28.6) | 1,071 (30.1) | 4,936 (28.3) |
| Obese, n (%) | 1,198 (50.3) | 5,453 (41.1) | 838 (44.4) | 3,647 (40.0) | 1,505 (42.3) | 7,306 (41.9) |
| BA.4/BA.5 +, n (%) | 400 (16.8) | 2,315 (17.4) | 187 (9.9) | 1,369 (15.8) | x | x |
| Up-to-date vaccination, n (%) | 622 (26.1) | 4,378 (33.0) | 519 (27.5) | 2,978 (34.4) | 922 (25.9) | 5,736 (32.9) |
| Unadjusted PF (95% CI), % | 5.5 (-2.8-13.2) |  | 38.0 (29.9-43.3) |  | x |  |
| Adjusted PF (95% CI), %, (p-value) * | 11.9(0.8-21.8) (p=0.036) |  | 45.9(36.2-54.1) (p<0.0001) |  | x |  |

\*Adjusted for age, gender, and comorbidities. Both exposures (previous SARS-CoV-2 infection and vaccination) were included in the model.

### Protection against reinfection with Omicron subvariants BA.4/BA.5 by prior SARS-CoV-2 infection

In unadjusted analyses, previous infection with BA.1/BA.2 provided modest protection against reinfection during the period of BA.4/BA.5 predominance with a PF of 38.0% (95% CI, 29.9-43.3%). However, prior Delta infections did not provide any protection against BA.4/BA.5 reinfection in unadjusted analyses. PF of BA.4/BA.5 infections by prior Delta infection was 5.5% (−2.8 to 13.2%).

Protection against reinfection stratified by age is shown in Table 2. Older adults (>65 years) infected during BA.1/BA.2 appeared to derive greater protection against reinfection than did younger adults (<65).

**Table 2.** Protection against Reinfection with the Omicron subvariants BA.4/BA.5 by age group

| Wave | Age group (years) | BA.4/BA.5 reinfection cases, N |  | BA.4/BA.5 reinfection incidence proportion, % |  | Preventable fraction, % | Pairwise P-value |
| --- | --- | --- | --- | --- | --- | --- | --- |
|  |  | Exposed | Unexposed | Exposed | Unexposed |  |  |
| Delta | 18-64 | 419 | 1,865 | 19.6% | 19.4% | -1.1% | <0.001 |
|  | ≥65 | 102 | 1,241 | 11.7% | 17.0% | 30.9% | Ref |
| BA.1/BA.2 | 18-64 | 192 | 1,035 | 12.4% | 17.9% | 30.9% | <0.001 |
|  | ≥65 | 60 | 792 | 6.9% | 15.6% | 55.6% | Ref |

**Table 3.** BA.4/BA.5 infection outcomes by prior COVID Status and preventable fraction (PF) of hospitalizations

|  | Delta+<br>(n= 400) | Delta-<br>(n= 2,315) | BA1BA2+<br>(n = 187) | BA1BA2-<br>(n=1,369) |
| --- | --- | --- | --- | --- |
| Hospitalization, n(%) | 164 (41.1) | 963 (41.6) | 47 (25.1) | 628 (45.9) |
| ICU admission, n(%) | 11 (2.8) | 93 (4.0) | 5 (2.7) | 62 (4.5) |
| Mechanical ventilation, n(%) | 5 (1.3) | 19 (0.8) | 0 | 15 (1.1) |
| Hospitalization, Unadjusted PF (95% CI), % | 1.3 (-9.9 – 12.0) |  | 44.6 (31.7-55.7) |  |
| Hospitalization, Adjusted PF (95%CI), % (p-value)* | 10.7% (95%CI, 4.9-21.7) (p=0.003) |  | 18.8% (95%CI, 10.3-28.3) (p<0.0001) |  |

\*Adjusted for age, gender, and comorbidities. Both exposures (previous SARS-CoV-2 infection and vaccination) were included in the model.

After adjusting for age, gender, BMI, comorbidities (HTN, DM, CKD, CHF, stroke), and up-to-date vaccination status, prior infection with BA.1/BA.2 still provided modest protection against BA.4/BA.5 reinfection. Prior BA.1/BA.2 infection conferred 45.9% (95% CI, 36.2-54.1)(p<0.0001) protection against reinfection with BA.4/BA.5. After adjustment, prior Delta infection provided no protection against BA.4/BA.5, 11.9% (95%CI, 0.8-21.8); p=0.036. In the Delta model, up-to-date vaccination status provided significant protection against BA.4/BA.5, with a PF of 22.7% (95%CI, 14.8-29.8); p<0.0001. The efficacy of up-to-date vaccination against BA.4/BA.5 in the BA.1/BA.2 model was 29.8% (95%CI, 20.5-38.1); p<0.0001.

### Prevention of hospitalization

In the unadjusted analysis, previous infection with SARS-CoV-2 during the BA.1/BA.2 wave offered 44.6% (95% CI, 31.7-55.7) protection against hospitalization during the BA.4/BA.5 wave. Delta infections did not confer significant protection against hospitalization during BA.4/BA.5. However, after adjusting for age, gender, comorbidities, and vaccination status, prior Delta infection was effective at preventing 10.7% (95%CI, 4.9-21.7)(p=0.003) of BA.4/BA.5 hospitalizations, while BA.1/BA.2 infection prevented 18.8% (95%CI, 10.3-28.3); p<0.0001. Efficacy of up-to-date vaccination in preventing hospitalization, after adjustment, was 20.0% (95%CI, 13.9-25.7)(p<0.0001) in the Delta model, and 22.9% (95%CI, 15.5-29.6)(p<0.0001) in the BA.1/BA.2 model.

## Discussion

In this retrospective cohort study of 20,987 individuals tested for SARS-CoV-2, previous Delta infection provided no protection against reinfection with Omicron subvariants BA.4/BA.5 and minimal protection against hospitalization. However, prior infection with earlier Omicron subvariants BA.1/BA.2 offered significant, albeit modest, protection against both reinfection and hospitalization. Up-to-date vaccination provided modest protection against hospitalization for both patients included in the Delta and BA.1/BA.2 analyses. Unexpectedly, we found that protection against reinfection was greater for older adults than for those under the age of 65 years.

Our study demonstrates that infection with pre-Omicron variant Delta offers no significant protection against BA.4/BA.5 infection. This is consistent with *in vitro* studies showing complete immune evasion of BA.4/BA.5 by neutralizing antibodies induced by prior infections or vaccination. Clinical studies have shown a progressive decline in protection conferred by prior SARS-CoV-2 infection against newer variants. While SARS-CoV-2 infection provided 80-90% protection against reinfection with pre-Omicron variants [2-9], studies of the initial Omicron variant found that past infection, at least with Delta, offered some protection, but not at the levels seen with earlier variants [10,16]. A recent study in Qatar found that pre-Omicron infections were 27.7% (95% CI, 19.3 to 35.2) effective at preventing BA.4/BA.5 infections [15]. It is important to note that they matched cases and controls by the week of their COVID test, which could better adjust for potential waning in natural immunity over time. However, we have previously shown that waning does not occur for at least 12 months [4,6]. Additionally, their study included individuals who received antigen testing and adjusted for testing indication. Their population was generally young and asymptomatic, and they therefore did not report on hospitalizations. Our cohort included an older, less healthy population, who received PCR testing in a healthcare setting rather than home antigen testing. Our cohort may therefore represent a population at higher risk of SARS-CoV-2 reinfection or adverse outcomes. Together, their findings and ours highlight a dramatic decline in the efficacy of natural immunity against newer SARS-CoV-2 variants, which may be most profound for our vulnerable populations.

Surprisingly, we found that following infection with BA.1/BA.2, older adults derived greater protection against BA.4/BA.5 than did younger adults. Prior studies have shown that natural immunity against COVID declines with advancing age. These unexpected findings, also seen with BA.1 [10], may be explained by differences in social behavior. It is plausible that following infection with COVID, older adults perceive their vulnerability and take greater precautions to avoid reinfection. Conversely, younger adults may feel they are protected against reinfection and no longer worry about catching it. Therefore they take fewer precautions, putting them at higher risk of exposure and infection. However, no differences were seen between younger and older adults with prior Delta infections. This suggests that the results were not completely a behavioral artifact. One explanation is that infection is dose dependent. Behaviors of older patients may have reduced the size of the inoculum and because the patients infected with BA.1/BA.2 had some immunity, they avoided infection, whereas older patients who had been infected with Delta were susceptible even to a very small exposure. Alternatively, the impact of infection on behavior may have been short-lived.

Prior infection with BA.1/BA.2, and to a minimal degree, Delta, also offered significant protection against hospitalization. Rates of ICU admission and mechanical ventilation were remarkably low during BA.4/BA.5, at only 4.0 and 0.9%, respectively, making it impossible to assess the impact of prior infection on these more serious outcomes.

Lastly, these findings are a sobering reminder of the challenges in preventing the spread of COVID-19, and the need for ongoing vaccine development. Our findings suggest that natural immunity is insufficient to prevent reinfections with immunologically different variants. While in vitro studies suggested that vaccination may be ineffective against BA.4/BA.5 [17], we found that up-to-date vaccination was modestly effective at preventing BA.4/BA.5 reinfection and hospitalization, even for patients without prior exposure to Omicron. These findings provide some reassurance and emphasize the importance of recent vaccination. Ongoing development of variant specific vaccines is needed to further improve their protective effect.

The strengths of this study include its large sample size, validity of PCR-confirmed SARS-CoV-2 infection, and adjustment for vaccine status, age, and common comorbidities. Inclusion of these demographic and clinical characteristics allowed us to explore the separate roles of natural immunity and vaccination in preventing SARS-CoV-2 reinfections, as well as their interactions.

While we were able to adjust for certain demographic variables and comorbidities, this study has several important limitations. Due to the rapid uptake of home antigen testing following the Delta wave, there may have been unmeasured differences between patients who presented for testing in the clinical setting versus testing at home. Additionally, greater time elapsed between Delta infections and BA.4/BA.5 exposure, than for BA.1/BA.2 infections. Several studies have shown that natural immunity persists for at least one year before waning [4,6]. However, our study alone cannot exclude the possibility that the lack of protection conferred by Delta infection is due to waning immunity. This study is also unable to account for differences in patient behavior involving avoidance of SARS-CoV-2 exposures. While age may account for some differences in behavior, we cannot further differentiate natural immunity from avoidance of exposures. Still, the extent to which we found minimal impact of previous infection remains concerning.

In conclusion, previous infection with Delta conferred no protection against reinfection with BA.4/BA.5 and minimal protection against hospitalization, regardless of vaccine status or age. BA.1/BA.2 infection offered limited protection against both. It seems unlikely that herd immunity will be possible. Future vaccines are necessary if more severe variants begin to circulate. Such vaccines will require rapid dissemination, prior to the emergence of new variants, in order to be effective as a public health intervention.

## Data Availability

All data produced in the present study are available upon reasonable request to the authors

## Notes

### Competing Interest Statement

The authors have declared no competing interest.

### Funding Statement

This study did not receive any funding

### Author Declarations

The Cleveland Clinic Institutional Review Board approved this work.

